# Association of Gene Variant Type and Location with Breast Cancer Risk in the General Population

**DOI:** 10.1101/2024.10.11.24315237

**Authors:** Mwangala P. Akamandisa, Nicholas J. Boddicker, Siddhartha Yadav, Chunling Hu, Steven N. Hart, Christine Ambrosone, Hoda Anton-Culver, Paul L. Auer, Clara Bodelon, Elizabeth S. Burnside, Fei Chen, Heather A. Eliassen, David E. Goldgar, Christopher Haiman, James M. Hodge, Hongyan Huang, Esther M. John, Rachid Karam, James V. Lacey, Sara Lindstroem, Elana Martinez, Jie Na, Susan L. Neuhausen, Katie M. O’Brien, Janet E. Olson, Tuya Pal, Julie R. Palmer, Alpa V. Patel, Tina Pesaran, Eric C. Polley, Marcy E. Richardson, Kathryn Ruddy, Dale P. Sandler, Lauren R. Teras, Amy Trentham-Dietz, Celine M. Vachon, Clarice Weinberg, Stacey J. Winham, Song Yao, Gary Zirpoli, Peter Kraft, Jeffrey N. Weitzel, Susan M. Domchek, Fergus J. Couch, Katherine L. Nathanson

## Abstract

**Importance:** Pathogenic variants (PVs) in *ATM, BRCA1, BRCA2, CHEK2*, and *PALB2* are associated with increased breast cancer risk. However, it is unknown whether breast cancer risk differs by PV type or location in carriers ascertained from the general population.

**Objective:** To evaluate breast cancer risks associated with PV type and location in *ATM, BRCA1, BRCA2, CHEK2*, and *PALB2*.

**Design:** Age adjusted case-control association analysis for all participants, subsets of PV carriers, and women with no breast cancer family history in population-based and clinical testing cohorts.

**Setting:** Twelve US population-based studies within the Cancer Risk Estimates Related to Susceptibility (CARRIERS) Consortium, and breast cancer cases from the UK-Biobank and an Ambry Genetics clinical testing cohort.

**Participants:** 32,247 women with and 32,544 age-matched women without a breast cancer diagnosis from CARRIERS; 237 and 1351 women with *BRCA2* PVs and breast cancer from the UKBB and Ambry Genetics, respectively.

**Exposure(s):** PVs in ATM, BRCA1, BRCA2, CHEK2, and PALB2.

**Main Outcome(s) and Measure(s):** PVs were grouped by type and location within genes and assessed for risks of breast cancer (odds ratios (OR), 95% confidence intervals (CI), and p-values) using logistic regression. Mean ages at diagnosis were compared using linear regression.

**Results:** Compared to women carrying *BRCA2* exon 11 protein truncating variants (PTVs) in the CARRIERS population-based study, women with *BRCA2* ex13-27 PTVs (OR=2.7, 95%CI 1.1-7.9) and ex1-10 PTVs (OR=1.6, 95%CI 0.8-3.5) had higher breast cancer risks, lower rates of ER-negative breast cancer (ex13-27 OR=0.5, 95%CI 0.2-0.9; ex1-10 OR=0.5, 95%CI 0.1-1.0), and earlier age of breast cancer diagnosis (ex13-27 5.5 years, p<0.001; ex1-10 2.4 years, p=0.17). These associations with ER-negative breast cancer and age replicated in a high-risk clinical cohort and the population-based UK Biobank cohort. No differences in risk or age at diagnosis by gene region were observed for PTVs in other predisposition genes.

**Conclusions and Relevance:** Population-based and clinical high-risk cohorts establish that PTVs in exon 11 of *BRCA2* are associated with reduced risk of breast cancer, later age at diagnosis, and greater risk of ER-negative disease. These differential risks may improve individualized risk prediction and clinical management for women carrying *BRCA2* PTVs.

**Key Points:** *Question:* Does *ATM*, *BRCA1*, *BRCA2*, *CHEK2* and *PALB2* pathogenic variant type and location influence breast cancer risk in population-based studies?

*Findings:* Breast cancer risk and estrogen receptor status differ based on the type and location of pathogenic variants in *BRCA2*. Women carrying protein truncating variants in exon 11 have a lower breast cancer risk in the population-based cohorts, older age at diagnosis and higher rates of estrogen receptor negative breast cancer than women with exon 1-10 or exon 13-27 truncation variants in population-based and clinical testing cohorts.

*Meaning:* Incorporating pathogenic variant type and location in cancer risk models may improve individualized risk prediction.

## Introduction

Pathogenic variants (PVs) in *ATM, BRCA1, BRCA2, CHEK2,* and *PALB2* are associated with increased risk for breast cancer and other cancer types^1–11^. PV carrier status informs clinical management decisions regarding surveillance, as with mammography and/or magnetic resonance imaging (MRI), risk reducing mastectomies, and treatment following a cancer diagnosis, as tumors may be sensitive to platinums and poly-ADP-ribose polymerase (PARP) inhibitors^12–16^.

Multiple intrinsic and extrinsic factors, such as polygenic risk scores (PRS), reproductive factors, environmental exposures, and variant type and location within the gene, have been demonstrated to be associated with differential breast cancer risk in women who carry PVs in high and moderate penetrance breast cancer susceptibility genes^17–19^. In high risk families, protein truncating variants (PTVs) in *BRCA1* exon 11 are associated with reduced breast cancer risk and increased ovarian cancer risk relative to PTVs in other parts of the gene^20–23^. Thus, exon 11 has been termed the ovarian cancer cluster region (OCCR). Similarly, exon 11 PTVs in *BRCA2* have been associated with higher ovarian cancer risk (*BRCA2* OCCR), and lower breast cancer risk compared to non-exon 11 PTVs^22–25^. A recent high-risk family-based study found that in women over age 50 years, *BRCA1* missense PVs were associated with lower breast cancer risk relative to PTVs^26^. Lower but non-significantly different breast cancer risk was identified for *BRCA2* missense PV carriage^26^. These studies in high-risk cohorts consistently have demonstrated that PV location, and, to a lesser extent, PV type influences the magnitude of breast cancer risk in *BRCA1* and *BRCA2* PV carriers.

PV type in moderate penetrance genes also has been evaluated as associated with differential risk of breast cancer. In *ATM*, studies in high-risk breast cancer cohorts have been inconsistent, with some demonstrating similar risks for breast cancer for PTVs and missense PV carriers, and others suggesting that rare missense PVs in the FAT and kinase domains confer higher breast cancer risk than PTVs^27–29^. A dominant negative missense variant, *ATM* c.7271T>G (p.V2424G), has been shown to increase breast cancer risk more than PTVs or other missense PVs^27,28,30,31^. Several association studies have suggested that *CHEK2* missense PVs confer a lower breast cancer risk than *CHEK2* PTVs^1,2,32–34^. Presently, only PTVs in *PALB2* have been associated with increased breast cancer risk; no differences in cancer risk by PV location have been demonstrated^9,34,35^.

To date, one population-based study has evaluated the breast cancer risk associated with *ATM, BRCA1, BRCA2, CHEK2,* and *PALB2* PTVs or missense PVs; but differential risk associated with other PV types or with PV location was not evaluated. Missense PVs in all five genes were associated with lower risk than PTVs or no risk^2^. A follow-up study evaluated the breast cancer risk associated with rare missense PVs in these genes and found a small fraction of missense PVs with breast cancer risk comparable to PTVs in *BRCA1*, *BRCA2*, and *ATM*; lower or no evidence of risk were suggested for *CHEK2* and *PALB2* missense PVs, respectively^34^. Herein we analyze breast cancer risk by PV type, functional effect, and location in *ATM, BRCA1, BRCA2, CHEK2,* and *PALB2* for the first time in population-based studies.

## Methods

Details of the CARRIERS consortium, overall study design, and participant characteristics have been reported previously^1^. CARRIERS follows the Strengthening the Reporting of Observational Studies in Epidemiology reporting guidelines^36^. For primary analyses, we included participants from 12 population-based studies within the CARRIERS consortium. Validation studies for specific *BRCA2* variant types were performed in women from the population-based United Kingdom Biobank (UKBB) and a high-risk Ambry Genetics clinical testing cohort (Supplemental Methods) ^37^. Informed consent was obtained from all participants; studies were approved by relevant ethical review boards.

### Variant Classification

All variants and classifications are included in Supplement eTable 1. Variants were classified as described previously^22^; details are in Supplemental Methods. Briefly, PVs included were 1) PTVs, accounting for the last known PTV in the gene; 2) large genomic rearrangements; and 3) missense, splicing or in-frame deletion (IFD) variants annotated as pathogenic or likely pathogenic by at least two commercial testing laboratories in ClinVar or by the Consortium of Investigators of Modifiers of BRCA1/2^22^. Variants were then annotated by type and functional interpretation (nonsense mediated decay [NMD], no NMD, IFD, missense, NMD/re-initiation) NMD is the mechanism by which transcripts with premature termination codons are degraded^38^.

A pre-planned analysis was performed that evaluated breast cancer risk associated with 1) PV location; 2) PV type and functional interpretation; 3) recurrent PVs (more than 20 instances in CARRIERS); and 4) whether the *BRCA2* or *PALB2* PV has been reported in patients with Fanconi Anemia (FA) ^39^.

### Statistical Analysis

For each gene separately, logistic regression was used to assess the association between variant type or location and risk of breast cancer by estimating odds ratios (OR) and 95% confidence intervals. Within each gene, subset analyses were performed to assess risk of breast cancer for pre-planned subgroups of PVs; the reference group was 1) frameshift for variant type; 2) NMD for functional interpretation; and 3) gene specific exons for location. Additional analyses by gene were stratified by first-degree family history of breast cancer. For *BRCA2*, predefined analysis of risk associations by ER-status and variant location (inside or outside exon 11) were performed using logistic regression. All analyses were adjusted for age at diagnosis for cases and at enrollment for age-matched unaffected controls. Linear regression was used to compare the means of age at diagnosis. For all analyses, two-sided p-values were calculated.

## Results

In 12 population-based CARRIERS studies comprising 32,247 women with and 32,544 without a breast cancer diagnosis, we evaluated differential breast cancer associations by PV type and location in *ATM, BRCA1, BRCA2, CHEK2,* and *PALB2*. We excluded 234 cases and 153 controls as they carried PVs in other established breast cancer genes, leaving 32,013 and 32,391 women with and without breast cancer for analysis. The mean (SD) ages at diagnosis in cases and enrollment in controls were 62.0 (11.5) and 61.2 (11.8) years, respectively. The PV prevalence and self-reported ancestry of the study participants is shown in Table 1 and eTable 2.

**Table 1:** Characteristics of population-based study participants.

|  | **Case**<br>**N=32247 %** |  | **Control**<br>**N=32544 %** |  | **Total**<br>**N=64791 %** |  |
| --- | --- | --- | --- | --- | --- | --- |
| **Study** |  |  |  |  |  |  |
| BWHS | 1437 | 4.5 | 2896 | 8.9 | 4333 | 6.7 |
| CPS3 | 1537 | 4.8 | 1729 | 5.3 | 3266 | 5.0 |
| CPSII | 4037 | 12.5 | 3935 | 12.1 | 7972 | 12.3 |
| CTS | 2226 | 6.9 | 2134 | 6.6 | 4360 | 6.7 |
| MCBCS | 4517 | 14.0 | 3249 | 10.0 | 7766 | 12.0 |
| MEC | 3641 | 11.3 | 3689 | 11.3 | 7330 | 11.3 |
| MMHS | 291 | 0.9 | 1257 | 3.9 | 1548 | 2.4 |
| NHS | 2088 | 6.5 | 2420 | 7.4 | 4508 | 7.0 |
| NHS2 | 935 | 2.9 | 1391 | 4.3 | 2326 | 3.6 |
| WCHS | 2215 | 6.9 | 1705 | 5.2 | 3920 | 6.1 |
| WHI | 4994 | 15.5 | 4535 | 13.9 | 9529 | 14.7 |
| WWHS | 4329 | 13.4 | 3604 | 11.1 | 7933 | 12.2 |
| **Pathogenic variants** |  |  |  |  |  |  |
| *ATM* | 253 | 0.8 | 139 | 0.4 | 392 | 0.6 |
| *BRCA1* | 273 | 0.8 | 35 | 0.1 | 308 | 0.5 |
| *BRCA2* | 421 | 1.3 | 84 | 0.3 | 505 | 0.8 |
| *CHEK2* | 351 | 1.1 | 139 | 0.4 | 490 | 0.8 |
| *PALB2* | 148 | 0.5 | 38 | 0.1 | 186 | 0.3 |
| **Pathogenic variants in family history negative participants** |  |  |  |  |  |  |
| *ATM* | 183 | 0.6 | 104 | 0.3 | 287 | 0.4 |
| *BRCA1* | 149 | 0.5 | 13 | 0.0 | 162 | 0.3 |
| *BRCA2* | 253 | 0.8 | 58 | 0.2 | 311 | 0.5 |
| *CHEK2* | 236 | 0.7 | 109 | 0.3 | 343 | 0.5 |
| *PALB2* | 86 | 0.3 | 31 | 0.1 | 117 | 0.2 |
| **Race or ethnic group** |  |  |  |  |  |  |
| N-Missing | 179 |  | 46 |  | 225 |  |
| Asian | 1282 | 4.0 | 1269 | 3.9 | 2551 | 3.9 |
| African American | 3946 | 12.3 | 4954 | 15.2 | 8900 | 13.7 |
| Hispanic | 1019 | 3.2 | 998 | 3.1 | 2017 | 3.1 |
| Non-Hispanic White | 25287 | 78.9 | 24770 | 76.2 | 50057 | 77.3 |
| Other | 534 | 1.7 | 507 | 1.6 | 1041 | 1.6 |
| **Age** |  |  |  |  |  |  |
| Mean (SD) | 62.0 (11.5) |  | 61.2 (11.8) |  | 61.6 (11.6) |  |
| Range | 21.0 - 94.0 |  | 21.8 - 94.3 |  | 21.0 - 94.3 |  |

## Risk by Variant Location

We evaluated breast cancer risk stratified by PV location. Relative to PTVs in exon 11, PTVs in *BRCA2* exons 1-10 were associated with a significantly increased breast cancer risk (OR=2.7, 95%CI 1.1-7.9, p=0.048; Figure 1a; eTable 3). PTVs in exons 13-27 also were associated with increased risk although not statistically significant (OR=1.6, 95%CI 0.8-3.5; Figure 1a; eTable 3). *BRCA2* exon 13-27 PTV carriers were diagnosed with breast cancer 5.5 years earlier on average than exon 11 PTV carriers (p<0.001; Figure 1a); carriers of PTVs in exons 1-10 also had a lower, albeit non-significantly different, age at diagnosis. Carriers of PVs predicted to not lead to NMD (non-NMD, e.g., missense, in-frame deletions [IFD], PTVs with stop codons in the last exon) had a non-significantly higher age at diagnosis relative to exon 11 PTV carriers (Figure 1a; eTable 3).

**Figure 1:**
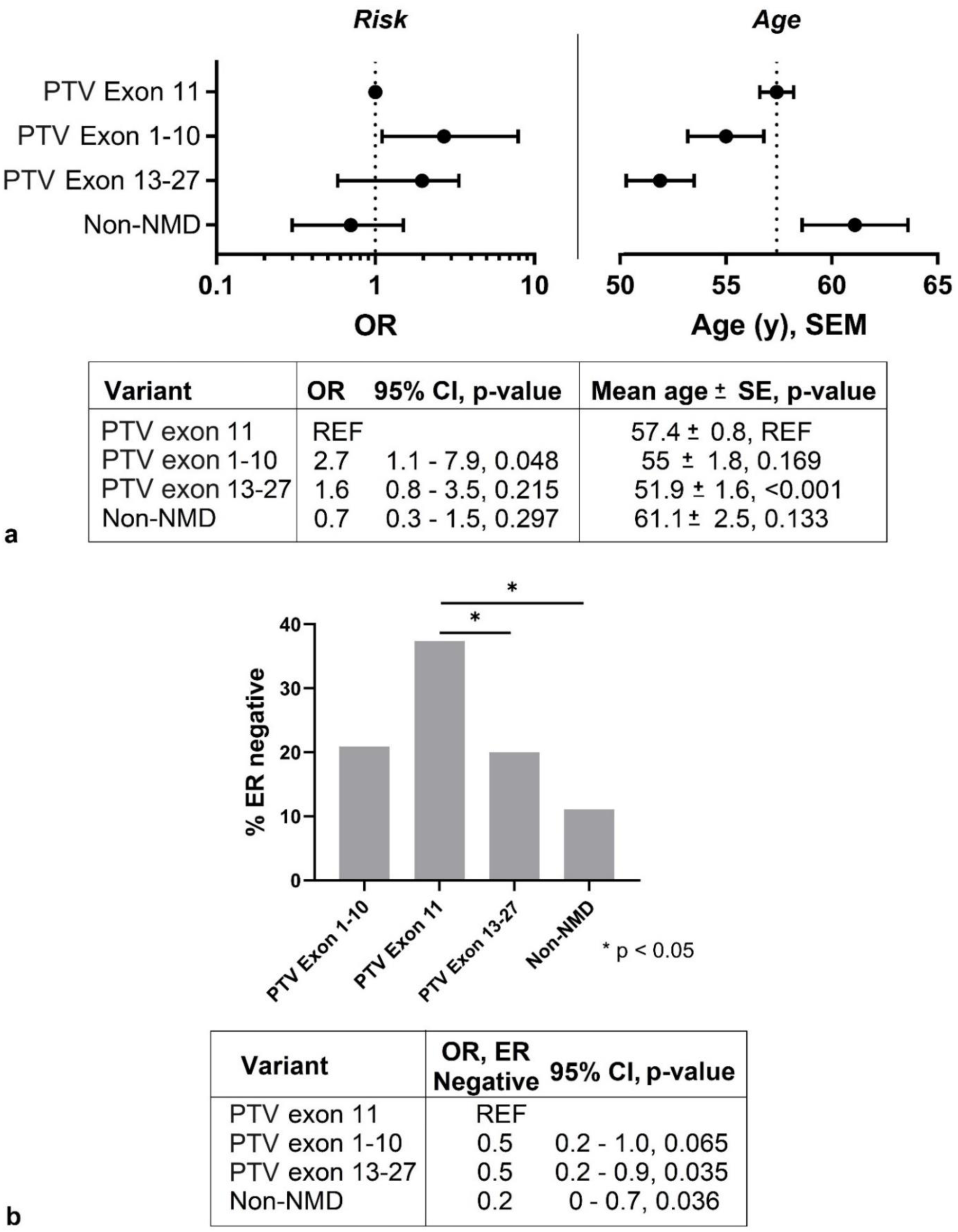
Age adjusted risk, age at breast cancer diagnosis, and ER-status in *BRCA2* variant carriers by PTV location and variant type. a. Top left, odds ratios (OR) and 95% confidence intervals (CI) in carriers of *BRCA2* pathogenic variants (PV). Dotted line is OR = 1. Top right, mean age at breast cancer diagnosis ± standard error of mean (SEM). Dotted line is 57.4 years. b. Estrogen-receptor (ER) expression in *BRCA2* PV carriers. ORs were estimated using logistic regression. PTV = protein truncating variant. Non-NMD includes in-frame deletions, missense PVs, and C-terminal PTVs not leading to NMD.

Approximately 70% of *BRCA2* PV carriers have estrogen receptor (ER)-positive breast cancer ^40,41^. We evaluated whether ER-status varied among women in CARRIERS with *BRCA2* PVs stratified by location and functional interpretation. Carriers of PTVs in exons 13-27 (OR=0.5, 95%CI 0.2-0.9, p=0.035) and non-NMD PVs (OR=0.2, 95%CI 0.0-0.7, p=0.036) had significantly lower rates of ER-negative breast cancer than carriers of exon 11 PVs (Figure 1b; eTable 4). Exon 1-10 PTV carriers also had lower, but not statistically significant, rates of ER-negative disease.

We sought to replicate these findings using the UKBB population-based study and a clinical testing cohort. Consistent with the findings from CARRIERS, women carrying exon 13-27 PTVs were diagnosed with breast cancer at significantly younger ages within the clinical testing cohort (44.9 ± 0.8 vs 47.1 ± 0.4, p=0.007) and UKBB (50.8 ± 1.5 vs 54.0 ± 0.8, p=0.031) than those with exon 11 PTVs (Figure 2a). Similarly, as within CARRIERS, women from the clinical testing cohort with PTVs in *BRCA2* exons 13-27 (OR=0.6, 95%CI 0.4-0.8, p = 0.006) and exons 1-10 (OR 0.5, 95%CI 0.4-0.8, p=0.003) had significantly lower rates of ER-negative breast cancer compared to those with exon 11 PTVs (Figure 2b; eTable 5). There was no difference in ER-status or age at diagnosis between exon 11 PTVs and non-NMD PVs in the clinical testing cohort.

**Figure 2:**
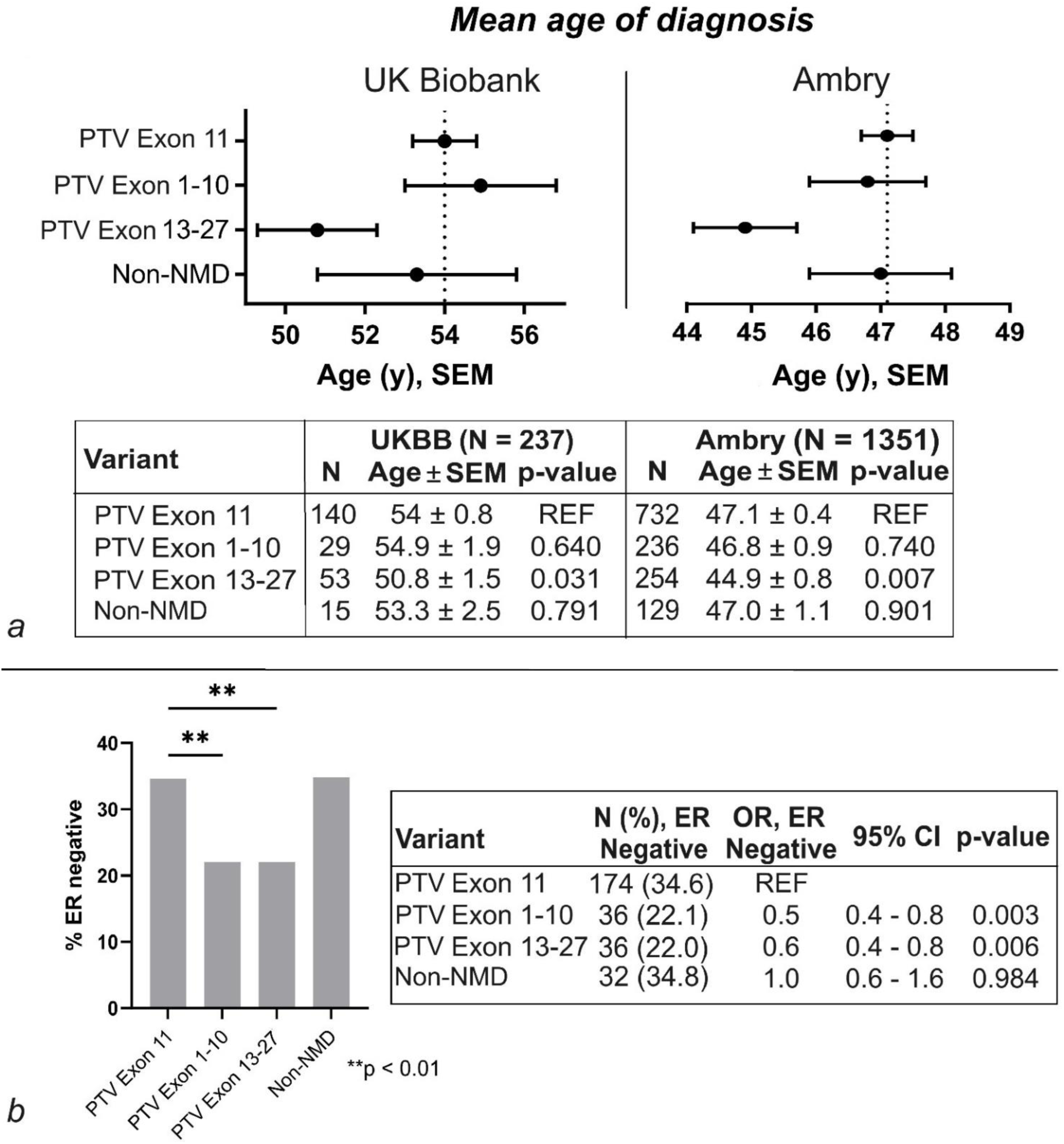
Mean age at diagnosis and association with ER-negative breast cancer in the United Kingdom Biobank population-based and Ambry Genetics clinical testing cohorts. a. Mean age at first breast cancer (BrCa) diagnosis by pathogenic variant (PV) location/type in women from the United Kingdom (UK) biobank and Ambry genetics. SEM = standard error of the mean. b. Prevalence and association of *BRCA2* PV location/type with estrogen receptor (ER)-negative BrCa in women from Ambry genetics. ORs were estimated using logistic regression PTV = protein truncating variant. Non-NMD includes in-frame deletions, missense PVs, and C-terminal PTVs not leading to NMD. Dotted line is OR = 1. CI = confidence interval.

We did not find an association between PV location and breast cancer risk when comparing *BRCA1* coding exon 10 (referred to as exon 11 in high-risk studies) PTV carriers to exons 1-9 or exons 11-23 PTV carriers, or carriers of *PALB2* PVs within the WD40 domain (exons 6-13) to those outside (Figure 3; eTables 6, 7). We did not evaluate association by location in *ATM* or *CHEK2* as PVs were not evenly distributed across the gene.

**Figure 3:**
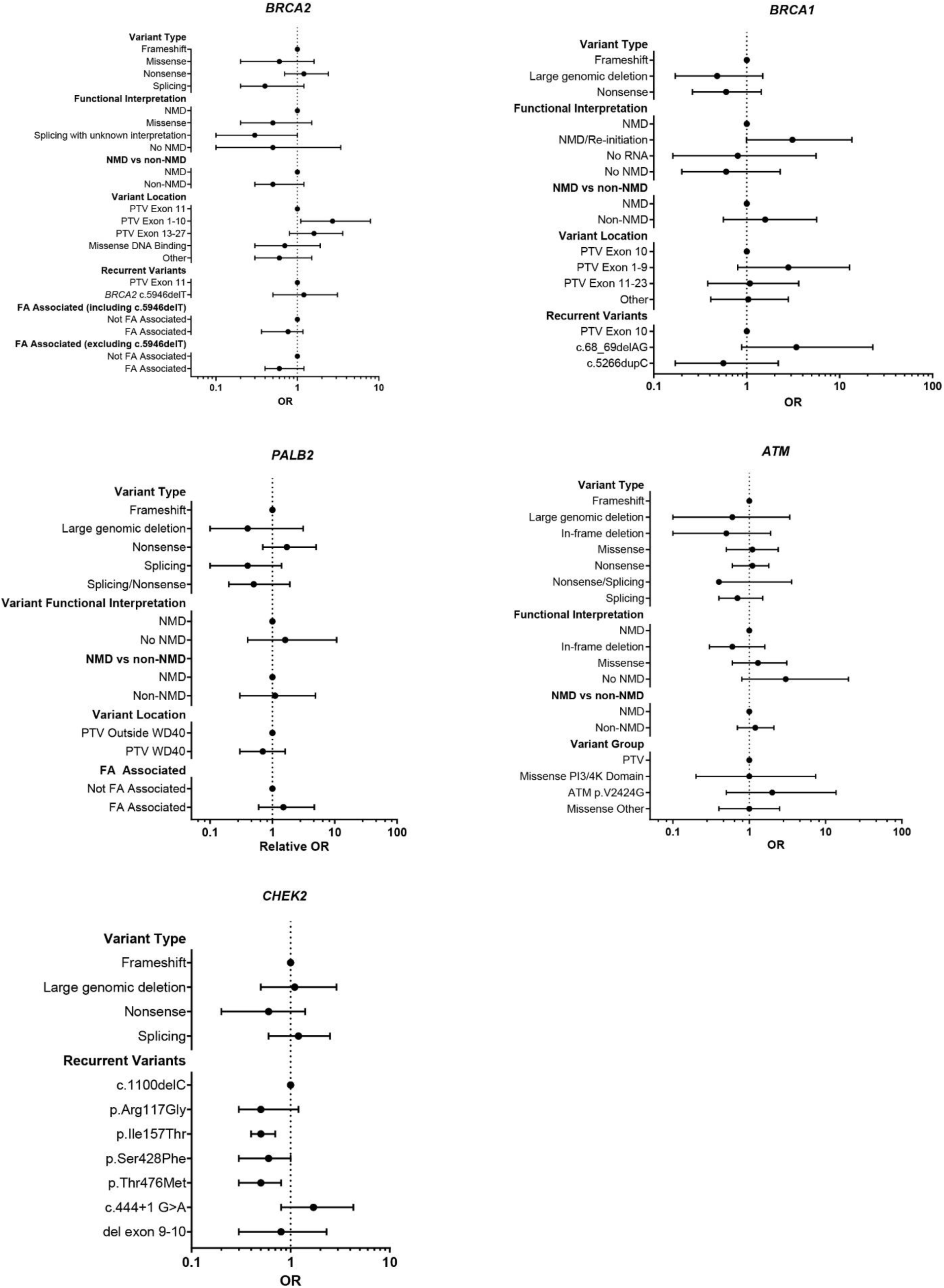
Age adjusted risk of *ATM*, *BRCA1*, *BRCA2*, *CHEK2*, and *PALB2* variant types and locations. Odds ratios (OR) and 95% confidence intervals (CI) were estimated using logistic regression in carriers of *BRCA2, BRCA1, PALB2*, *ATM*, and *CHEK2* pathogenic variants (PVs) in the population-based studies. PTV = protein truncating variant. Non-NMD includes in-frame deletions, missense PVs, and C-terminal PTVs not leading to NMD. Dotted line is OR = 1.

## Risk by Variant Type

We evaluated the breast cancer associations of *ATM, BRCA1, BRCA2, CHEK2* and *PALB2* PVs stratified by PV type, functional interpretation, and by variant frequency. We found no significant difference in breast cancer risk comparing *ATM, BRCA2,* or *PALB2* PVs by variant type, functional interpretation, or FA-association in all population-based study participants, and those with no breast cancer family history (Figure 3; eTables 3,7,8,9,10). Although the breast cancer risk associated with non-NMD PVs in *BRCA2* was not significantly different than that of PTVs resulting in NMD, the data suggested a lower risk (OR=0.5, 95%CI 0.3-1.2, p=0.118; Figure 3; eTable 3). The breast cancer risk associated with *ATM* p.V2424G was elevated but not significantly different from the association for all PTVs (OR=2.0, 95%CI 0.5-13.7, p=0.378; Figure 3; eTable 10).

In *BRCA1,* PTVs upstream of p.Met128 are predicted to result in re-initiation of translation at p.Met128 after degradation of the variant transcript by NMD^42–44^. Women in CARRIERS with PTVs predicted to lead to NMD/re-initiation (pre-Met128) may have a higher breast cancer risk compared to carriers of PTVs after Met128 (OR=3.1, 95%CI 1.0-13.5, p=0.083), as observed in high-risk families^22^ (Figure 3; eTable 6). Although the risk of breast cancer associated with c.5266dupC eastern European founder variant was not significantly different from that associated with coding exon 10 PTVs, the data were suggestive of a relatively lower risk, as seen in high-risk families^22^. We could not evaluate the difference in breast cancer risk between women with *BRCA1* PTVs and missense PVs as missense PVs were only observed in breast cancer cases and not controls. In women without a family history of breast cancer, risk was significantly lower for carriers of *BRCA1* large genomic deletions (OR=0.1, 95%CI 0.02-0.86, p=0.028) and nonsense PVs (OR=0.2, 95%CI 0.04-0.85, p=0.034) as compared to frameshift PV carriers (Figure 4; eTable 11).

**Figure 4:**
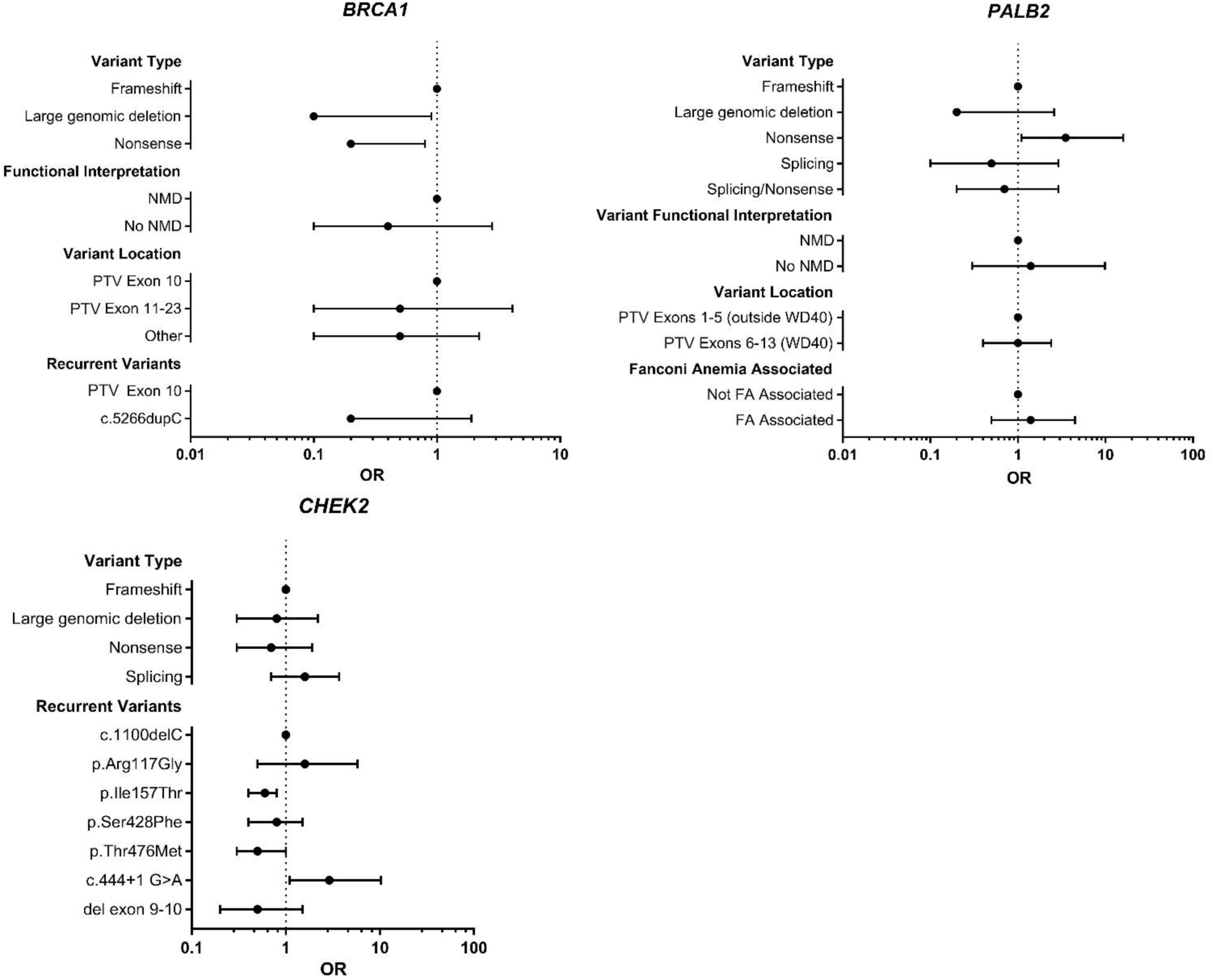
Age adjusted risk of *BRCA1*, *CHEK2* and *PALB2* variant types and locations in family history negative participants. Odds ratios (OR) and 95% confidence intervals (CI) were estimated using logistic regression in carriers of *BRCA1*, *CHEK2*, and *PALB2* pathogenic variants (PV) from participants with no family history of breast cancer in first degree relatives. PTV = protein truncating variant. Non-NMD includes in-frame deletions, missense PVs, and C-terminal PTVs not leading to NMD. Dotted line is OR = 1.

We compared the breast cancer risk associated with the recurrent *CHEK2* PVs c.349A>G (p.Arg117Gly), c.470T>C (p.Ile157Thr), c.1283C>T (p.Ser428Phe), c.1427C>T (p.Thr476Met), c.444+1G>A, and del exon 9-10 to that of c.1100delC. Carriers of p.Ile157Thr (OR= 0.5, 95%CI 0.4 - 0.7, p< 0.001) and p.Thr476Met (OR=0.5, 95%CI 0.3-0.8, p=0.005) had a significantly lower breast cancer risk than c.1100delC carriers (Figure 3; eTable 12). Carriers of the founder splicing variant c.444+1G>A had the highest breast cancer risk of all *CHEK2* recurrent variants compared to noncarriers (OR=4.2, 95%CI 1.9-10.5, p< 0.001), however, it was non-significantly different from that of c.1100delC carriers (OR=1.7, 95%CI 0.8-4.3, p=0.230). In women without a family history of breast cancer, the breast cancer risk was significantly higher in c.444+1G>A carriers than c.1100delC carriers (OR=2.9, 95%CI 1.1-10.3, p=0.054; Figure 4; eTable 13).

## Discussion

In the population-based CARRIERS study, we evaluated whether *ATM, BRCA1, BRCA2, CHEK2*, and *PALB2* PVs, stratified by type, functional interpretation, and location, were associated with breast cancer risk in a pre-planned analysis. Many of the genotype associations with differences in breast cancer risk observed in high-risk populations also were found in this population-based cohort at similar odds ratios, although several did not reach statistical significance, likely due to small numbers of PV carriers. Within *BRCA2*, we found that variant location for PTVs and functional effect (non-NMD) was associated with significant differences in breast cancer risk, age of diagnosis, and ER-status; the latter two have not been previously evaluated in high-risk cohorts. The results support the importance of considering not only the gene in which the PV resides, but variant type and location as modifiers of effects in breast cancer risk modeling with downstream implications for counseling patients.

Prior studies in high-risk cohorts have demonstrated an approximately two-fold increase in breast cancer risk for women with PTVs in regions before (5’) and after (3’) *BRCA2* exon 11 compared to PTVs in exon 11^22,23,45^. We similarly found an increased risk in carriers of *BRCA2* PTVs outside of exon 11; carriers of PTVs 5’ of exon 11 had an almost three-fold significantly increased risk, and 3’ of exon 11 had risk elevated 1.6-fold, albeit not at statistical significance. Prior studies suggested that the lower risk of breast cancer associated with PTVs in exon 11 is specific to the c.5946delT founder PTV. However, we found no difference in risk between c.5946delT and other exon 11 PTVs; the lower risk relative to PTVs outside exon11 was consistent across exon 11^46^. We also found that carriers of *BRCA2* PTVs within exons 13-27 have a significantly younger mean age at diagnosis than carriers of PTVs in exon 11 (51.9 vs 57.4), a finding that replicated in women within both the clinical testing (44.9 vs 47.1) and population-based UKBB (50.8 vs 54.0) cohorts despite differing mean ages at diagnosis across the studies. Women with non-NMD PVs in CARRIERS had a later (61.1), although not significantly different, mean age at diagnosis relative to women with exon 11 PVs (57.4). Thus, depending upon the PV type and location, breast cancer risk associated with PVs in *BRCA2* may vary up to four-fold with consequent differences in the age at diagnosis-with PTVs outside of exon 11 having the higher risk and youngest age at diagnosis and non-NMD variants the lowest risk and oldest age at diagnosis, with breast cancer risk and age at diagnosis associated with exon 11 PTVs falling in the middle.

Within CARRIERS, we observed higher rates of ER-negative breast cancer in women with *BRCA2* exon 11 PTVs as compared to carriers of PTVs after and before exon 11, although non-significantly in the latter. Similarly, in the clinical testing dataset, women with exon 11 PTVs had higher rates of ER-negative breast cancer than women with PTVs outside exon 11. The relatively higher rates of ER-negative breast cancer among *BRCA2* exon 11 PTV carriers, with an older age at diagnosis, agrees with findings from a previous family-based cohort that found higher rates of ER-negative breast cancer in *BRCA2* PV carriers over age 50 years^47^. These data suggest that *BRCA2* exon 11 PTVs may be associated with a relatively increased risk of ER-negative breast cancer leading to an ‘ER-negative cluster region’, similar to the OCCR for ovarian cancer and possibly a cluster region for pancreatic cancer^22,24,45,48^. Ovarian cancer and ER-negative breast cancer have multiple genomic similarities including high rates of *TP53* PVs and PI3-kinase pathway dysregulation^49–53^. Although the *BRCA2* OCCR has been recognized for over 20 years, the biological mechanism underlying the relatively increased risk of ovarian cancer has not been determined. At 4932 base pairs long, *BRCA2* exon 11 is one of the largest internal exons in the human exome^54^. The efficiency of NMD is significantly reduced in long exons and directly related to the distance between the PTV and downstream exon junction^55^. An early study evaluated NMD for eight PTVs in *BRCA2* exon 11 lymphoblastoid cell lines (LCLs), but has multiple limitations; absolute levels of difference in expression and statistical significance was not reported^56^. Further, the efficiency of NMD varies by tissue and cell type so the degree of NMD in LCLs may not be representative of breast tissue^38^.

Additionally, truncated proteins have been reported in cell lines and breast tissue in studies of *BRCA2* exon 11 PTVs, suggesting that for at least some PTVs, NMD does not occur^57,58^. Thus, variation in NMD efficiency may contribute to the differing phenotypes observed in carriers of exon 11 PVs. Our study supports the need for further functional studies to elucidate the biological basis of the differential risks for ovarian cancer and ER-negative breast cancer based on PV location in *BRCA2*.

We did not observe any differences by PTV location in *BRCA1,* likely due to small numbers. However, we did observe a higher risk of breast cancer for *BRCA1* PTVs with NMD/re-initiation as compared to other PTVs, consistent with high-risk cohorts^20–22^.

Larger sample sizes will be needed to determine type and location associations in *BRCA1* PV carriers in the general population. We found that carriers of *CHEK2* p.Ile157Thr and p.Thr476Met had significantly lower breast cancer risk than c.1100delC carriers, as previously reported^2,33,59–61^. The c.444+1A>G variant was associated with the highest breast cancer risk among recurrent *CHEK2* PVs particularly in women with no breast cancer family history. These data are consistent with a prior study of *CHEK2* PV carriers, identified through clinical testing^59,62^.

We did not find differences in *ATM* breast cancer association by variant type, differing from studies that have suggested higher risks for *ATM* missense variants over PTVs^27,28^. Although individual rare missense variants may confer higher risk, the breast cancer risk of missense PVs in aggregate did not differ from that of PTVs. We found no significant differences in breast cancer risk by PV type or location in *PALB2* PV carriers.

The major limitation of this study is that despite including 64,791 women, the numbers of individuals with PVs in each gene were small, limiting the statistical power.

Consequently, although several of our findings are consistent with findings in prior high-risk cohorts, they did not reach statistical significance. Future studies with larger populations should improve statistical power and further refine the risk estimates.

In summary, our findings confirm the heterogeneity of risk depending upon by PV type and location in breast cancer susceptibility genes in a population-based cohort. The findings are consistent with high-risk cohorts and expand upon them with considerations of age of diagnosis and ER-status. Currently, breast cancer risk models, such as CanRisk and Tyrer-Cuzick, do not include variant type and PV location within genes as determinants of risk^63,64^. The results of this study suggest this information should be included as a component of breast cancer risk models for carriers of PVs in *BRCA1, BRCA2*, and *CHEK2*, given their notable impact on risk. These findings support the importance of considering predisposition gene variant type and location, as they differentially contribute to breast cancer risk and thus may impact the clinical management of PV carriers^65,66^.

## Data Availability

All data produced in the present study are available upon reasonable request to the authors.

https://www.ncbi.nlm.nih.gov/projects/gap/cgi-bin/study.cgi?study_id=phs002820.v1.p1

## Acknowledgements

BWHS: Breast cancer pathology data were obtained from several state cancer registries, including some or all of the following: AZ, CA, CO, CT, DE, DC, FL, GA, IL, IN, KY, LA, MD, MA, MI, NJ, NY, NC, OK, PA, SC, TN, TX, VA. The content of the manuscript is solely the responsibility of the authors and does not necessarily represent the official views of the National Cancer Institute, the National Institutes of Health, or the state cancer registries. The IRBs of participating institutions and cancer registries have approved this research, as required. The authors thank participants and staff of the Black Women’s Health Study for their contributions.

CPSII: The authors express sincere appreciation to all Cancer Prevention Study-II and Cancer Prevention Study-3 participants, and to each member of the study and biospecimen management group. The authors would like to acknowledge the contribution to this study from central cancer registries supported through the Centers for Disease Control and Prevention’s National Program of Cancer Registries and cancer registries supported by the National Cancer Institute’s Surveillance Epidemiology and End Results Program.

NHS and NHS2: The study protocol was approved by the institutional review boards of the Brigham and Women’s Hospital and Harvard T.H. Chan School of Public Health, and those of participating registries as required. We would like to thank the participants and staff of the NHS and NHS2 for their valuable contributions as well as the following state cancer registries for their help: AL, AZ, AR, CA, CO, CT, DE, FL, GA, ID, IL, IN, IA, KY, LA, ME, MD, MA, MI, NE, NH, NJ, NY, NC, ND, OH, OK, OR, PA, RI, SC, TN, TX, VA, WA, WY. The authors assume full responsibility for analyses and interpretation of these data.

WHI: Program Office: (National Heart, Lung, and Blood Institute, Bethesda, Maryland) Jacques Rossouw, Shari Ludlam, Joan McGowan, Leslie Ford, and Nancy Geller.

Clinical Coordinating Center: (Fred Hutchinson Cancer Research Center, Seattle, WA) Garnet Anderson, Ross Prentice, Andrea LaCroix, and Charles Kooperberg. Investigators and Academic Centers: (Brigham and Women’s Hospital, Harvard Medical School, Boston, MA) JoAnn E. Manson; (MedStar Health Research Institute/Howard University, Washington, DC) Barbara V. Howard; (Stanford Prevention Research Center, Stanford, CA) Marcia L. Stefanick; (The Ohio State University, Columbus, OH) Rebecca Jackson; (University of Arizona, Tucson/Phoenix, AZ) Cynthia A. Thomson; (University at Buffalo, Buffalo, NY) Jean Wactawski-Wende; (University of Florida, Gainesville/Jacksonville, FL) Marian Limacher; (University of Iowa, Iowa City/Davenport, IA) Jennifer Robinson; (University of Pittsburgh, Pittsburgh, PA) Lewis Kuller; (Wake Forest University School of Medicine, Winston-Salem, NC) Sally Shumaker; (University of Nevada, Reno, NV) Robert Brunner Women’s Health Initiative Memory Study: (Wake Forest University School of Medicine, Winston-Salem, NC) Mark Espeland.

This research has been conducted using data from UK Biobank, a major biomedical database www.ukbiobank.ac.uk.

Conflict of Interest: E.C.P has received research funding from Grail, A.V.P has received institutional funding from Grail, K.R. received research funding from Medtronic and reports family and institution have filed intellectual property for the application of artificial intelligence for diagnosis and risk stratification. J.E.O. has research funding from Exact Sciences, S.M.D. received honoraria from AstraZeneca and GlaxoSmithKline, and institutional research funding from AstraZeneca and Clovis Oncology. J.N.W. was employed by and holds stock options with Natera, consulted for Myriad Genetics, and served on AstraZeneca’s speaker’s bureau. K.L.N. consulted for Merck, P.K. received travel and accommodation expenses from Regeneron. S.Y. received institutional research funding from Repare Therapeutics and AstraZeneca. F.J.C. received honoraria from Ambry Genetics/Konica Minolta, US Oncology Network, Natera, consulted for AstraZeneca, served on speaker’s bureaus for Ambry Genetics and Qiagen, and received research funding and travel accommodations and expenses from Grail. R.K., M.E.R., and T.P. are employed by Ambry Genetics.

## Abbreviations of Study Names

BWHS: The Black Women’s Health Study
CPS3: Cancer Prevention Study-3 Cohort
CPSII: Cancer Prevention Study-II Nutrition Cohort
CTS: California Teachers’ Study
MCBCS: Mayo Clinic Breast Cancer Study
MEC: Multiethnic Cohort Study
MMHS: Mayo Mammography Health Study
NHS: The Nurses’ Health Study
NHS2: The Nurses’ Health Study 2
WCHS: Women’s Circle of Health Study
WHI: The Women’s Health Initiative
WWHS: Wisconsin Women’s Health Study

## Supplemental Methods

### Variant Characterization

Variants were described using the Human Genome Variation Society nomenclature with nucleotide numbering starting from the A of the ATG initiation codon. Each variant was individually reviewed by KLN, with sub-set of variants in addition reviewed by a larger group (MPA, NJB, CH, FJC, SNH). PV type was designated as the DNA-level changes (frameshift, missense, nonsense, splicing, large genomic deletions or duplications). The functional consequence was predicted for each PV as 1) leading to nonsense mediated decay (NMD); 2) not leading to NMD (no NMD); 3) in-frame deletion; 4) resulting in no RNA; 5) missense; 6) unknown; or 7) NMD/re-initiation (*BRCA1* only). PTVs with the stop codons before the last 50 base pairs of the penultimate exon in all genes were predicted to result in NMD, otherwise their functional interpretation was no NMD. The no RNA functional interpretation was assigned to deletions that included the initiation codon in the first exon. For *BRCA1*, PTVs occurring before Methionine128 were predicted to result in NMD with re-initiation. We only included missense variants located in putative functional domains using the boundaries defined in the Pfam database that were annotated as pathogenic in Clinvar by at least two commercial testing laboratories^1^. We defined FA associated variants using those included in published work^2^.

To evaluate the PV location, we subset PTV (nonsense, frameshift, and splicing) PVs into those in exon 10 (annotated as exon 11 in prior studies), before (exon 1-9), and after (exon 11-23) in *BRCA1* PV carriers and PTVs in exon 11, exon 1-10, and exon 13-27 in *BRCA2*. *BRCA2* exon 12 did not have any PTVs. In *PALB2*, we compared carriers of PTVs within the WD40 domain to those outside using Pfam defined boundaries for the WD40 domain (exons 6-13).

### Ambry Genetics Clinical Testing Cohort

The study was approved by the Western Institutional Review Board (WIRB) (now WCG), which exempted review of the clinical testing cohort. The clinical testing cohort consisted of a nationwide sample of 9,887 with DCIS who underwent clinically indicated hereditary cancer multigene panel testing at Ambry Genetics from March 2012 to December 2016. Data on patient characteristics were collected from requisition forms at the time of testing and clinical notes provided by ordering clinicians. For comparison, information on 38,057 women with IDC who underwent germline genetic testing during the same time period at Ambry Genetics were extracted. The diagnoses and tumor pathology for 10% of the DCIS and invasive disease patients were confirmed in medical records as part of a validation study. Women who had previously undergone testing for *BRCA1* or *BRCA2* prior to multigene panel testing were excluded.

### UKBB Population-Based Cohort

Participants for the replication of *BRCA2* findings were ascertained from the United Kingdom Biobank (UKBB), a population-based cohort study with 470,000 participants with whole exome sequencing agnostic of diagnosis (UKBB project 69929)^3^. Sequencing data are linked to electronic health records and the UK National Cancer Registration and Analysis Service (NCRAS). Participants were included in our replication cohort if they carried *BRCA2* PVs and a diagnosis of breast cancer in the linked cancer register data. 237 women met inclusion criteria. Age was abstracted from cancer register as well; ER-status was not available. The UKBB is approved by the UK Biobank Research Ethics Committee with informed consent from all participants.

